# Can a vaccine-led approach end NSW’s outbreak in 100 days, or at least substantially reduce morbidity and mortality?

**DOI:** 10.1101/2021.08.18.21262252

**Authors:** Laxman Bablani, Tim Wilson, Hassan Andrabi, Vijaya Sundararajan, Driss Ait Ouakrim, Patrick Abraham, Jason Thompson, Tony Blakely

**Affiliations:** Centre for Epidemiology and Biostatistics, Melbourne School of Population and Global Health, University of Melbourne, Melbourne, Australia; Department of Medicine, University of Melbourne, Melbourne, Australia; Department of Public Health, La Trobe University, Melbourne, Australia; Health Economics Unit, Centre for Health Policy; Melbourne School of Population and Global Health; University of Melbourne; Transport, Health and Urban Design Research Lab; Melbourne School of Design; University of Melbourne

## Abstract

**Background and Aims:** The New South Wales (NSW) COVID-19 outbreak is at 478 daily cases on August 16, 2021.

Our aims were to:

1. estimate the time required to reach ≤5 cases per day under three lockdown strengths (weak, moderate, strong), and four vaccination rollouts: (a) per the original plan, (b) prioritizing essential workers, (c) b plus rapid vaccination of 25% of <60-year-olds with AstraZeneca (AZ25), and (d) b plus rapid vaccination of 50% of <60-year-olds with AstraZeneca (AZ50).
2. estimate the number of cases, hospitalizations, and deaths in the 100 days after 1/August for the 12 scenarios.

**Methods:** An agent-based model was adapted to NSW and the Delta variant. Hospitalization and mortality rates for unvaccinated COVID-19 infections were doubled given the virulence of Delta.

**Results:** The business-as-usual rollout fully vaccinates 50%, 70% and 80% of >16-year-olds by 10/Oct, 21/Nov, and 28/Dec, respectively. This reduced to 1/Oct, 30/Oct, and 22/Nov for the fastest (AZ50) rollout.

A strong lockdown with a rapid vaccine rollout was the fastest to reach ≤5 cases (14-day average), with a median of 78 days (90% Uncertainty interval 61 - 103) or 18/Oct, compared to 207 days (166 - 254) or 24/Feb for a weak lockdown with no rollout acceleration. Increased lockdown strength had more impact than rollout acceleration.

Under the AZ25 vaccination scenario, there were 1,440 (90% UI 262 - 10,600 deaths in the first 100 days of cases under a weak lockdown, compared to 71 (90% UI 26 - 178) under a strong lockdown scenario.

**Conclusion:** NSW will likely achieve 70% vaccination of >16-year-olds before reaching ≤5 daily cases. Accelerating the vaccine rollout is important for the medium-term, but in the short-term increased restriction strength was more effective at reducing caseload (and subsequently mortality and hospitalisation) than accelerating the vaccine rollout.

**Significance of Study**
“*The known*” NSW is facing a Delta-variant COVID-19 outbreak, with a vaccination-led strategy for controlling the outbreak. Despite the increased infectivity and virulence of the Delta-variant, little contemporaneous modelling exists. We, therefore, model several restrictions and vaccination scenarios.
“*The new*” NSW will likely achieve 70% vaccination of >16-year-olds before daily cases are ≤5. Increased lockdown strength was more effective at reducing cases than accelerating the vaccine rollout.
“*The implications*” Accelerating the vaccine rollout is important in the medium-term, but in the short-term strong public health and social restrictions (including lockdown) are more effective at reining in cases.

## Introduction

Australia currently has a policy goal of “zero community transmission of COVID-19”, which encourages early, sharp, lockdowns for new outbreaks as a key component of eliminating the virus. The recent roadmap approved by National Cabinet foresees a pivot to a strategy of increasingly “living with COVID-19”, particularly as 70% and 80% vaccination targets are met^1^.

The current NSW outbreak, starting June 16, was not eliminated early through a short, sharp lockdown – and as of 11 August has a five-day rolling average of more than 300 daily cases and a 4% per day average increase in cases over the preceding fortnight. The NSW Government did not immediately implement aggressive containment strategies akin to ‘stage 4’-style city-wide (or even state-wide) lockdowns as used elsewhere. Instead, the strategy appeared to be a geographically varying intensity of restrictions based on caseloads - the first state-wide lockdown was implemented at 6pm on 14 August 2021. This was combined with increasing vaccination rates, while targeting younger, critical workers. 50% of adult Sydneysiders were vaccinated with their first dose on 14 August 2021, which the NSW government contends will provide strategic space to ease restrictions on 28 August, even though most people under 40 years of age will, at that time, remain unvaccinated.

To assist policymaking in NSW, and public understanding, our objectives were:

1. To estimate how long it would take to reach fewer than five cases per day, under 12 combinations of three strengths of lockdown (weak, moderate, and strong) by four vaccination roll-out scenarios: (a) as per original plan, b) prioritizing essential workers with Pfizer, c) b plus rapid vaccination of 25% of <60-year-old adults with AstraZeneca, and d) b plus rapid vaccination of 50% of <60-year-old adults with AstraZeneca.
2. To estimate the number of cases, hospitalizations, and deaths in the 100 days after 1 August 2021 for the 12 above scenarios.

We used the COVID-19 Policy Model^2^, an agent-based model used to underpin Victoria’s exit from its second wave in 2020^3^ and used in peer-reviewed research on Victoria’s COVID-19 outbreak^4^. In applying this model to NSW and the latest outbreak, it was challenging to know exactly what levels of restrictions were being applied across Sydney and NSW, and how to capture all elements of heterogeneity that may matter (e.g., full heterogeneity of household size, essential workers, and social contact patterns in Western Sydney).

Thus, it is essential at the outset to emphasize that we are primarily attempting to forecast likely patterns of infection across the community, rather than predicting exact numbers of cases, hospitalisations, and deaths – although we do provide the latter by scenarios with wide uncertainty intervals. We use Monte Carlo simulations that include both stochastic (random) uncertainty and uncertainty in inputs, for example, uncertainty about vaccine efficacy.

## Methods

To reflect the increased infectivity of Delta we updated the model with a shorter time from infection to being infectious^5^ and increased peak infectivity^6^. We calibrated our model’s transmissibility profile parameters to achieve R0s of 5.5 to 6.5, and then draw from within this range for each run. We doubled the infection fatality ratios and hospitalisation rates we used for 2020 modelling given accruing evidence about Delta.^7^ See Supplementary Table 1 for further details of model parameterisation.

### Parameterising Sydney’s lockdown and vaccination strategies

The strictness of lockdown varies across NSW and varies over time. To eliminate COVID-19 in the community, the state needs to pull down the effective reproductive number (*R*_*eff*_), or the number of people a single case infects, to below 1 through vaccination, lockdowns, contact tracing, mask usage, or a combination of strategies. Despite NSW’s current restrictions, the rates are slowly rising (4% per day in the two weeks up to 10 August, equivalent to an *R*_*eff*_ of about 1.2).

The model uses pre-existing stage 3 (soft lockdown) and 4 (hard lockdown) restrictions calibrated for our previous Victorian modelling (Supplementary Table 2). Given the considerable variation in restriction settings across Sydney and NSW and uncertainty about how these restrictions compare to Victoria, we additionally specified a stage 3b (moderate lockdown) with parameters halfway between those of stages 3 and 4. Given the current *R*_*eff*_ in NSW, the moderate and soft lockdown scenarios (stage 3 and 3b) appear to bound what is currently playing out in NSW.

On top of these three lockdown strengths, the interventions we model are 1) a business-as-usual (BAU) vaccine roll-out per the current vaccine allocation horizons^8^ document, 2) shifting all Pfizer to essential workers and prioritising first doses, then back to BAU, 3) prioritizing essential workers as in 2, and also rapidly vaccinating 25% of adults aged under 60 years with AstraZeneca, and 4) as 3, increasing to 50% of adults aged under 60 years receiving AstraZeneca. The model contains the rates at which Pfizer and AstraZeneca become available, so switching cohorts to receive AstraZeneca increases the overall speed of the rollout – but also means that fewer young adults may receive Pfizer later in 2021 due to rapid use of AstraZeneca in August and September.

To quantify the health burden of each scenario, we calculated the number of hospitalisations and deaths caused by infections in the 100 days after 1 August. These rates do not allow – at very high caseloads – for any worsening case fatality rate with health system overload. We selected 100 days as it roughly approximated when the 70% or 80% adult vaccination target is achieved, noting this date varies with vaccination speed. In some scenarios, less than five cases per day occurred before 100 days – hospitalisation and deaths were only tallied up to that point. For simplicity, we estimate deaths and hospitalisations that occur among the cases in these 100 days; some of these hospitalisations and deaths will occur after 100 days due to lag-times.

## Results

Figure 1 shows the median daily cases over the preceding week for the four vaccination strategies, for each of the three-stage settings. For our first objective, the fastest option is Stage 4 with the most aggressive vaccination strategy, taking 78 days (90% uncertainty interval 61 to 103 days) from 1 August to reach an average of five daily cases over a 14 day period (or median of 18 Oct, 95% UI 1 Oct to 12 Nov). This time increases to 207 days (168 to 250 days) for a weak lockdown with no rollout acceleration (or 24 Feb 2022, 90% UI 16 Jan to 8 Apr).

**Figure 1:**
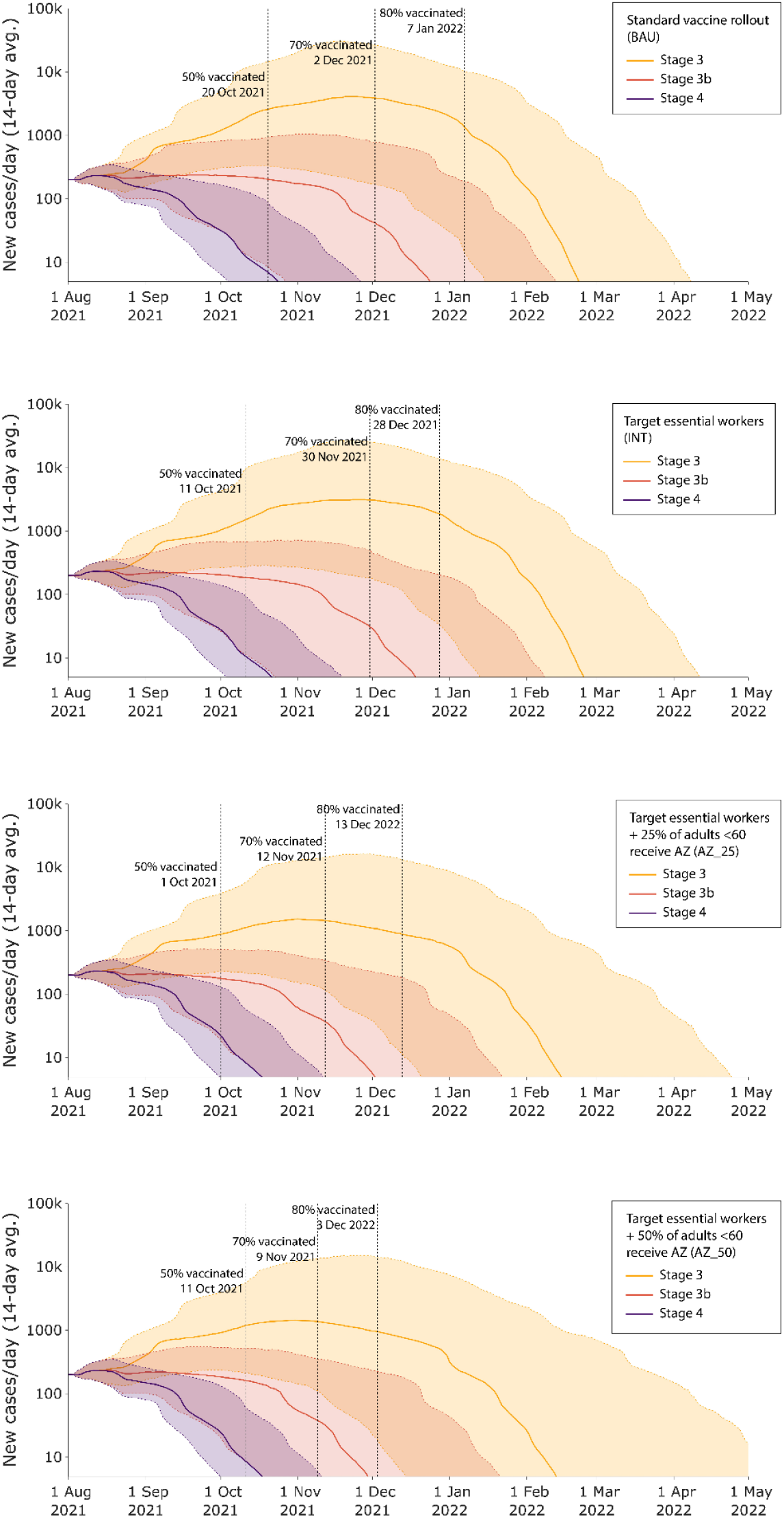
Median daily cases (log scale; 7-day average) for each combination of Stage and vaccination strategy. Vaccination thresholds include a 10-day period for the second vaccine dose to be fully effective. Solid lines are medians across 4000 runs of the ABM, and bands show the 5^th^ to 95^th^ percentile across runs.

For the more likely or realistic scenario of Stage 3b, and essential workers vaccinated first and 25% of < 60-year-olds opting to get AstraZeneca rapidly, it is 124 days (78 to 175) until fewer than 5 cases per day is achieved (3 Dec, 90% UI 18 Oct to 23 Jan).

Superimposed on Figure 1 are solid vertical lines for when 50%, 70% and 80% vaccination coverage of the population over 16-years-old is achieved for each vaccination strategy. Except for fewer than half of the Stage 4 lockdown runs, the date at which 50% of adults are double vaccinated (plus a 10-day buffer to allow immune response) occurs before less than five cases per day is achieved.

Figure 2 below shows heatmaps of the numbers of estimated cases, hospitalisations, and deaths for each vaccination strategy by stage combination, for a timeline of 100 days. Hospitalisations and deaths are given for infections acquired within the first 100 days, avoiding the lag between infections to hospitalisations and deaths, to make for clearer comparisons. Deaths from infections caused with a two-week lag from infections, reflecting infections in days 0 to 86, are in Supplementary Table 3.

**Figure 2:**
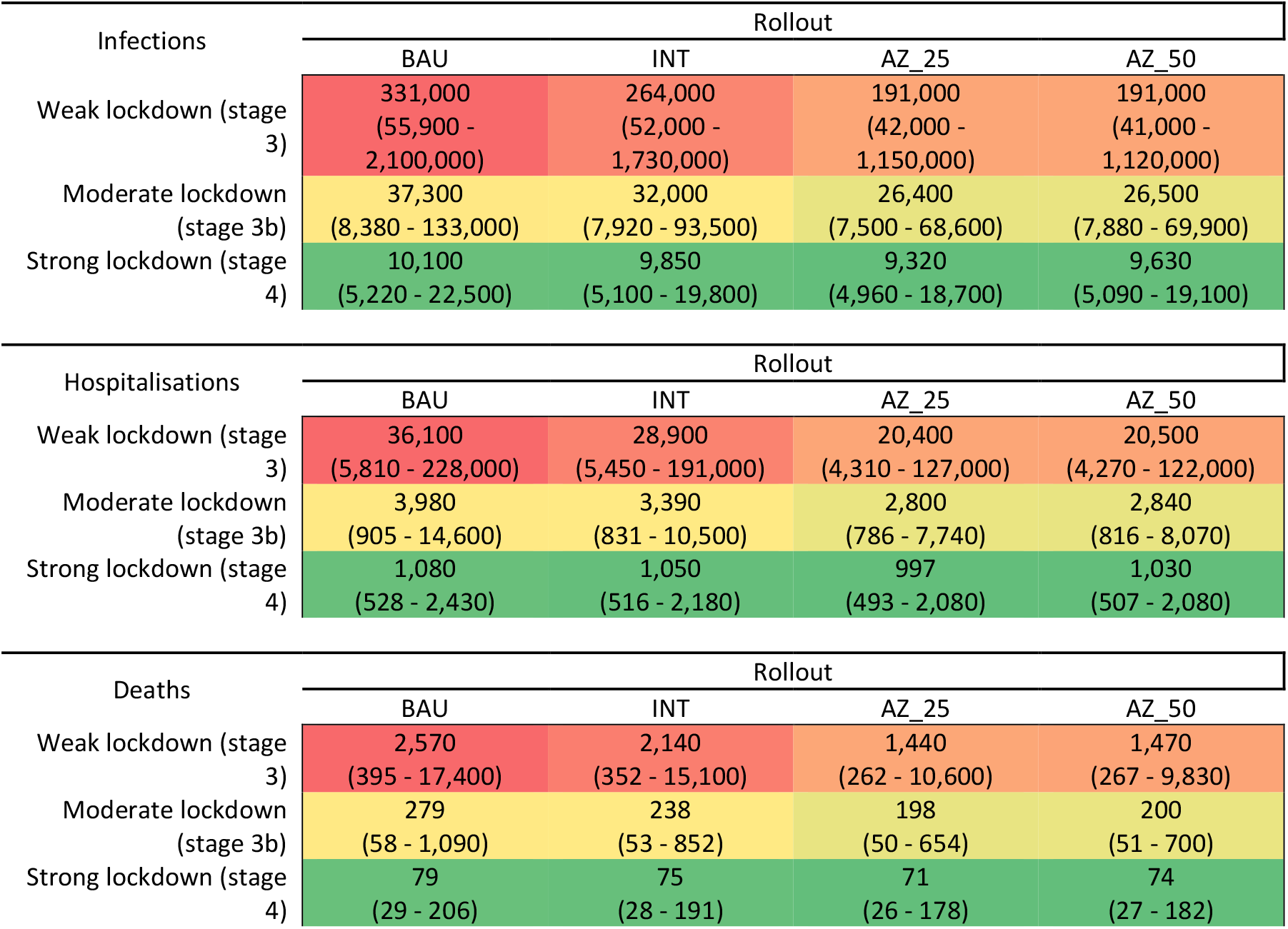
Heatmaps of cases, in the first 100 days of each strategy (or cases are less than five per day is achieved first). Hospitalizations and death are those among infections in first 100 days, and therefore the total hospitalizations are those up to (roughly) 10 days beyond 100 days due to time lags, and deaths are two to three weeks after infections to allow for time lags. BAU: vaccine roll-out per the National Plan; INT: shifting all Pfizer to essential workers as a priority then back to BAU; AZ_25: prioritizing essential workers as in INT and vaccinating 25% of adults aged under 60 years with AstraZeneca; AZ_50: increasing to 50% the adults aged under 60 years vaccinated with AstraZeneca.

The most substantial reduction in cases, hospitalisations and deaths is generated through tightening restrictions, i.e., going vertically down the columns in the heatmaps, rather than faster vaccination strategies presented horizontally, across the rows. For example, for deaths under the BAU vaccine rollout, median deaths increase 33-fold from 79 under strong lockdown to 2570 under weak lockdown. Under a strong lockdown, median deaths only increase by 7%, from 74 under a rapid vaccine rollout (AZ50) to 79 under a BAU vaccine rollout. However, there is an interaction, in that under a weak lockdown a slower BAU vaccine rollout with 2,570 deaths is 75% greater than the 1,470 deaths with a faster AZ50 rollout.

For what might be the most likely vaccine rollout scenario, AZ25, 1,440 (90% UI 262 - 10,600) deaths occur over 100 days under a weak lockdown, 198 (50 to 654) under a moderate lockdown, and 71 (90% UI 26 - 178) under a strong lockdown scenario. The number of infections and AZ25 for a weak lockdown are 191,000 (or an average of 1910 per day [not implausible given nearly 400 cases per day on August 13], 90% uncertainty interval 42,000 to 1,150,000), for a moderate lockdown 26,400 (7,500 to 68,600) and for a strong lockdown 9,320 (or an average of 93 cases per day [seemingly implausible now, unless there is a very strong turn-around], 4,960 to 18,700).

## Conclusion

NSW will likely achieve 70% vaccination of >16-year-olds before reaching fewer than five daily cases. Put another way, for most scenarios it took more than 100 days (and sometimes much longer) to ‘bend the curve’ and reach fewer than five cases per day. Accelerating the vaccine rollout is important for the medium-term in NSW (and the rest of Australia), as it will make the population more resilient to outbreaks later in 2021 and into 2022 when we open the borders. However, in the next few months, it is the strength of lockdown (or public health and social measures) that has the largest potential impact in reducing COVID-19 hospitalisations and deaths.

There were some modelling analyses performed on the Delta NSW outbreak in July 2021. The Burnet Institute^9^ analysed whether Victorian-style Stage 3 and Stage 4 restrictions would be sufficient to control the outbreak. They found it would take roughly 6 weeks to control the outbreak. Similarly, our previous modelling^10^ found that stage 4 restrictions would take around 5.8 weeks to get cases to five or less with longer times to exit restrictions for less stringent restrictions. However, at the time of publishing their study, restrictions in Sydney had recently been introduced and the impact was unknown. Also, since our previous modelling, we have updated the model for new data on Delta – the shift forward in time from infection to being infectious (which undermines in part the impact of contact tracing). Last, a nowcasting model^11^ found that the level of social distancing in NSW around 25 July 2021 was inadequate to control the outbreak and 80% of agents need to comply with distancing regulations to control the outbreak in a pandemic in a month. All studies, including this one, concur that strong restrictions would be required to turn around cases and head towards re-elimination. The modelling in this paper improves on previous efforts by updating the analyses to the observed scenario in August 2021. It also incorporates emerging evidence on the twicer higher virulence of the Delta variant ^7^.

Epidemiologically, a sustained and hard lockdown for (unfortunately) several months is necessary to minimize the heath loss due to COVID-19. The ‘best’ policy options require balancing the direct health impacts of SARS-CoV-2 with the unintended health impacts of sustained and hard lockdowns, and the social and economic impacts of lockdowns.

In applying our model to NSW, we want to emphasise it is only a model. It is becoming increasingly challenging to model Delta, especially amid changing and inconsistently applied public health interventions. Nevertheless, it seems most unlikely the conclusion that strong public health and social restrictions remain more important in the short term for constraining case numbers than vaccination, would be overturned by any plausible adjustments to the model.

## Data Availability

This was all publicly available data

---

**Supplementary Table 1:** Key parameter estimates and ‘agent’ characteristics most relevant to current paper used in the agent-based model

| Key Parameters | Parameter Estimate |
| --- | --- |
| **R0 (calibrated to via transmissibility in a situation with no interventions of any type.)** | Global uniform distribution 5.5 to 6.5. |
| **ChAdOx1 nCoV-19 (AstraZeneca) Vaccine effectiveness (VE) at reducing <u>any</u> infection (applies within Victoria only)** | Reduction in probability of AstraZeneca vaccinated contracting illness from unvaccinated infected varies depending on variant.<br>• Emary et al (2021)<sup>(Emary and et al 2021)</sup> report a VE of (66.5% (95% CI 37.1%, 82.1%) for alpha, and 80.7% (69.2%, 87.9%) for wild-type and other variants for all infections 14 days after the 2<sup>nd</sup> vaccine dose in an RCT in the UK.<br>• Pritchard et al (2021) undertook a large cohort study, also in the UK, estimating a VE of 79% after two doses (95% CI 65 to 88%; using OR in Suppl Table 6).<sup>(Pritchard, Matthews et al. 2021)</sup> The sample was split between Alpha (53%) and other variants (“S gene positive, plus 1 or 2 other genes”, 45%) – but did not include Delta variants (Suppl Table 3).<br>• Sheikh, et al (2021) report that AZ effectiveness for all infections against the Delta variant in Scotland was 60% (53–66%, S gene-positive).<sup>(Sheikh, McMenamin et al. 2021)</sup> Of note, against S gene-negative cases (Alpha) effectiveness was 73% (95% CI 66–78), which is compatible with results from Pritchard et al, and providing an estimate of the relative decrease in VE for Delta.<br>We parameterized ChAdOx1 nCoV-19 VE against any transmission to cover the estimates in above studies and allow for some deterioration with Delta: Beta: 40,26.66 (mean 61%, median = 61%, 2.5<sup>th</sup> %ile = 49%, 97.5<sup>th</sup> %ile = 72%) |
| **BNT162b2 (Pfizer) Vaccine effectiveness at reducing <u>any</u> infection (applies within Victoria only)** | Reduction in probability of Pfizer vaccinated contracting illness from unvaccinated infected also varies depending on variant.<br>• Polack, et al reported in their RCT predominantly set in the United States that vaccine efficacy was 95% effective (95% credible interval, 90.3 to 97.6) – but this did not include Delta.<sup>(Polack, Thomas et al. 2020)</sup><br>• Hass , et al in their nation-wide study from Israel reported a vaccine effectiveness of 95.3% (95% CI 94.9 to 95.7%) after 2 doses – but again not including Delta.<sup>(Haas, Angulo et al. 2021)</sup><br>• Pritchard et al (2021) estimated an effectiveness of (80%, 95% CI 74 to 85%; using OR in Suppl Table 6) after 2 vaccine dose in a sample evenly split between Alpha (53%) and other variants (but not including Delta, 45%) (Suppl Table 3).<sup>(Pritchard, Matthews et al. 2021)</sup><br>• Sheikh, et al (2021) report that AZ effectiveness for all infections against the Delta variant in Scotland was 79% (95% CI 75–82%).<sup>(Sheikh, McMenamin et al. 2021)</sup> Of note, against S gene-negative cases (Alpha, B.1.1.7) effectiveness was 92% (95% CI 90–93) – consistent with a lower VE for Delta. |
|  | We parameterized ChAdOx1 nCoV-19 VE against any transmission to cover the estimates in above studies and allow for some deterioration with Delta: Beta 35.55, 8.88 (mean = 80%, median = 81%, 2.5<sup>th</sup> %ile = 67%, 97.5<sup>th</sup> %ile = 90%). |
| **Vaccine and past infection effectiveness at reducing infectivity if infected**<br>**(applies to arriving cases [all assumed vaccinated] and within Victoria for infections among vaccinated)** | Harris et al found a 0.53 (95% CI 0.44 to 0.63) lower odds of infection among close household contacts of ChAdOx1 nCoV19 vaccinated infected (AZ) c.f. unvaccinated, and 0.49 (0.44 to 0.56) for BNT162b2 (Pfizer).<sup>(Harris, Hall et al. 2021)</sup> That is an average of 50% lower attack rate.<br>Such secondary attack rate studies for Delta are not available at the time of writing. However, there is concerning evidence that vaccinated people infected with Delta have the same virus levels as unvaccinated infected people<sup>12</sup>, implying that the above 50% lower attack rate may not apply.<br>Given this uncertainty, we parameterized the reduction in infectivity if vaccinated to range from 0% to 50% (Beta 1.9, 5.7; mean = 25%, median = 23%; 2.5<sup>th</sup> %ile = 3%, 97.5<sup>th</sup> %ile = 59%). |
| **Protection of past infection against new infection (including past infection wild-type protection against new variants)** | This is a highly uncertain variable, requiring estimating how much infection with current variants (e.g. Alpha, Delta) will protect individuals against infection from new variants in the future. So we specified a wide uncertainty. (Reinfection can only after 21 days.) Parameterization: Beta: 8, 2 (mean = 80%, median = 82%, 2.5<sup>th</sup> %ile = 52%, 97.5<sup>th</sup> %ile = 97%). |
| **ChAdOx1 to-19 (AstraZeneca) Vaccine effectiveness (VE) at reducing hospitalisation and death given infection** | We parameterized the VE for hospitalisation and death to both be consistent with the above studies, and coherent with the VE for any infection such that the expected value of VE for hospitalisation <u>conditional</u> on being infected was 50% (beta 50, 50), and the VE for death conditional on being infected was 70% (beta 58.8, 25.2). |
| **BNT162b2 (Pfizer) Vaccine effectiveness at reducing hospitalisation and death given infection** | We parameterized the VE for hospitalisation and death to both be consistent with the above studies, and coherent with the VE for any infection such that the expected value of VE for hospitalisation <u>conditional</u> on being infected was 50% (beta 50, 50), and the VE for death conditional on being infected was 70% (beta 58.8, 25.2). |
| **Waning vaccine and natural immunity** | Nil. Whilst there will likely be waning immunity over time, for the two-year duration of our modelling it may not be substantial. Second, we do not include booster vaccination – which we suspect is likely to occur within a year of initial vaccination, offsetting any waning immunity. |
| **Increased hospitalisation rate and mortality rate compared to that of Victoria in August 2020 due to Delta** | Ratio of 2.0 (95% UI 1.5 to 2.67; ln normal)<br>Recent studies show that Delta has approximately twice the hospitalisation and death rates of the previous variants<sup>7</sup>. |
| **Time post-infection to being infectious** | Infectiousness started on day 1 (the day after infection) at about a quarter of peak infectiousness, with peak infectiousness increasing linearly to time drawn (below), then falling linearly to the drawn value for the mean illness period (two panels below). |
| **Time to peak infectivity (days, log-normal)<sup>2</sup>(Bi, Wu et al. 2020)** | Per-agent log normal distribution: mean = 4.4, SD = 1.5 |
| **Mean illness period<sup>3</sup>** | Per-agent log normal distribution: mean = 21, SD = 2 |
| **Mean adherence with isolation of infected cases** (beta distribution) <sup>‡</sup> | Global beta distribution (beta 28, 2; median = 94.3%, SD = 4.5%) <sup>†</sup> |
| **Date of case simulation initialisation** (Day 0) | August 1<sup>st</sup>, 2021 |
| **Asymptomatic cases**<sup>3,4</sup> | Global normal distribution: mean = 30%, sd = 3%<br>Doubled for children. |
| **Infectiousness of asymptomatic cases vs symptomatic cases per contact**<sup>5</sup> | Global normal distribution: mean = 75%, sd = 3% |
| **Ratio transmission risk from mask wearing** | Global beta distribution (beta 40, 60).<br>Note this is per-person, so transmission is on average reduced by 69% ($1 - (0.4^2)$) if both are masked. |
| **Household size** | Scaled beta distribution with median 3.0. Beta 2.2, 2.2 scaled to [1, 5] with draws rounded to nearest integer. |
| **Ratio of transmissibility for asymptomatic versus symptomatic cases** | 0.75 [Source: https://www.cdc.gov/coronavirus/2019-ncov/hcp/planning-scenarios.html] (Applied to global transmissibility parameter.) |
| **Proportion of non-household infected contacts traced within the first three days, at a caseload of 5 per day.** | Beta distribution (2, 2) rescaled to [0.6, 0.8]<br>Mean 0.7 |
| **Chance of an infected non-household contact of a known case becoming known, per day.** | $\text{chance} = 1 / (\text{dailyCases} * ((a * \text{dailyCases}^{0.92} - b) + 50))$ $\text{dailyCases} := \text{daily cases averaged over the last week.}$<br>Solve for a and b with (dailyCases, chance) = (1, 0.53) and (5, c). Set c to attain the probability over 3 days drawn above. Note that (1, 0.53) was selected so that 90% of contacts are traced over three days at a caseload of one per day i. The following are typical single-day trace probabilities for the median draw (70% contacts traced after three days at 5 cases):<br>1, 54%; 2, 42%; 3, 37%; 5, 33%; 10, 28%; 50, 20%; 100, 16%; 500, 6.8%; 1000, 4.0%; 5000 0.95%.<br>This is designed to, on average, find no more than 50 cases per day via non-household contact tracing alone. |
| **Chance of an infected household contact of a known case becoming known, per day.** | 100% |
| **Proportion of undetected symptomatic cases who spontaneously reported themselves** | 0.5 |
| **Time of undetected symptomatic cases spontaneous self reporting** | 1-2 days after peak infectivity. |
| **Transmission multiplier of person who is complying with their isolation** | 0.33 |

Code for ABM at: https://github.com/population-interventions/CovidABM/tree/master/NSW
¥ Assumed parameter based on expert opinion in conjunctions with available public data sources such as Google COVID-19 mobility reports.

**Supplementary Table 2:** Specification of stages for weak (Stage 3), NSW moderate (Stage 3b), and strong (Stage 4) restrictions in the Agent Based Model

| Parameter | Description | Stage 3 | Stage 3b | Stage 4 |
| --- | --- | --- | --- | --- |
| Span | Mobility of the agents in the simulation, as a population level multiplier on top of underlying heterogeneity. | 5 | 4 | 3 |
| Proportion_People_Avoid | Proportion of agents that consider avoiding agents from other households. | 60% | 72.50% | 85% |
| Proportion_Time_Avoid | Proportion of time that agents who consider avoiding agents from other households actually do so. | 60% | 72.50% | 85% |
| Complacency_Bound | The two parameters above reduce at a rate of 1% per day in lockdown, down to the complacency bound. | 50% | 64% | 78% |
| Essential_Workers | Proportion of the workforce considered essential. Agents tagged as essential workers cannot avoid other agents or be avoided, and have higher Visit_Radius and Superspreader values. | 35% | 27.50% | 20% |
| SchoolsOpen | Whether schools are open, which causes students to act like essential workers. | No | No | No |
| Mask_Wearing | Proportion of agents that wear masks when outside their home. | 60% | 72.50% | 85% |
| Visit_Radius | Search radius for picking a random gathering location to visit. | 5 | 4.3 | 3.6 |
| Visit_Frequency | Probability per agent per day of visiting a nearby gather location, if any are within Visit_Radius. | 14.28% | 14.28% | 14.28% |
| Superspreaders | Probability per non-isolation compliant agent per day of visiting a random gather location. | 4% | 3% | 2% |

**Supplementary Table 3:** Heatmap of deaths in the first 100 days of each strategy, lagged by two weeks from infections.

| Deaths | Rollout |  |  |  |
| --- | --- | --- | --- | --- |
|  | BAU | INT | AZ_25 | AZ_50 |
| Weak lockdown (stage 3) | 1,688<br>(327 - 9,840) | 1,490<br>(293 - 8,498) | 1,088<br>(235 - 6,375) | 1,156<br>(241 - 6,178) |
| Moderate lockdown (stage 3b) | 254<br>(58 - 872) | 222<br>(53 - 725) | 191<br>(49 - 587) | 193<br>(51 - 632) |
| Strong lockdown (stage 4) | 79<br>(29 - 202) | 75<br>(28 - 188) | 71<br>(26 - 177) | 74<br>(27 - 181) |

